# How acceptable is rapid whole genome sequencing for infectious disease management in hospitals? Perspectives of those involved in managing nosocomial SARS-CoV-2

**DOI:** 10.1101/2022.06.15.22276423

**Authors:** Paul Flowers, Julie McLeod, Fiona Mapp, Oliver Stirrup, James Blackstone, Luke B Snell, Christine Peters, Emma Thomson, Alison Holmes, James Price, Dave Partridge, Laura Shallcross, Thushan I de Silva, Judith Breuer

## Abstract

**Background:** Whole genome sequencing (WGS) for managing healthcare associated infections (HCAIs) has developed considerably through experiences with SARS-CoV-2. We interviewed various healthcare professionals (HCPs) with direct experience of using WGS in hospitals (within the COG-UK Hospital Onset COVID-19 Infection (HOCI) study) to explore its acceptability and future use.

**Method:** An exploratory, cross-sectional, qualitative design employed semi-structured interviews with 39 diverse HCPs between December 2020 and June 2021. Participants were recruited from five sites within the larger clinical study of a novel genome sequencing reporting tool for SARS-CoV-2 (the HOCI study). All had experience, in their diverse roles, of using sequencing data to manage nosocomial SARS-CoV-2 infection. Deductive and inductive thematic analysis identified themes exploring aspects of the acceptability of sequencing.

**Findings:** The analysis highlighted the overall acceptability of rapid WGS for infectious disease using SARS-CoV-2 as a case study. Diverse professionals were largely very positive about its future use and believed that it could become a valuable and routine tool for managing HCAIs. We identified three key themes ‘1) ‘Proof of concept achieved’; 2) ‘Novel insights and implications’; and 3) ‘Challenges and demands’.

**Conclusion:** Our qualitative analysis, drawn from five diverse hospitals, shows the broad acceptability of rapid sequencing and its potential. Participants believed it could and should become an everyday technology capable of being embedded within typical hospital processes and systems. However, its future integration into existing healthcare systems will not be without challenges (e.g., resource, multi-level change) warranting further mixed methods research.

## Introduction

Whole genome sequencing (WGS) is a process of analysing DNA to identify genetic mutations and disease variants [1,2]. Since the first human genome was sequenced in 2000[3], WGS has been used for diagnosis and personalised medicine[^4]^ as well as epidemiological surveillance[5,6]. Following rapid advances in sequencing technology over the past two decades[3], WGS can identify between-patient transmission of infections ‘in real time’[7] and as such, can be used to assist with managing outbreaks and nosocomial infection[8]. During the COVID-19 pandemic[9,10,11], WGS was completed on the SARS-CoV-2 virus[12,13]. This allowed for insights into the evolution of the virus[14,9] and the identification of different variants[15,16,17].

A number of barriers to using WGS to assist with managing healthcare associated infections (HCAIs) have been identified including the high cost of implementation (e.g., attaining equipment and training staff)[18,8]; the lack of available protocols and guidelines, particularly targeting those new to its use[8,19]; the lack of infrastructure and capacity[8; the lack of bioinformatician availability and output interpretation[20,21]; challenges associated with complex software[20; ethical and legal concerns[22]; and uncertainty on whether it is best used as an IPC tool with a rapid turnaround time enabling agile responses, or its use across different outbreaks gradually providing retrospective insights at a slower pace.

Given the benefits and barriers of WGS for managing HCAIs, this current paper sought to understand the acceptability of using WGS for SARS-CoV-2 from a range of professional perspectives: infection prevention control (IPC) teams, other healthcare professionals (HCPs) such as bioinformaticians, those involved in diagnostic and sequencing laboratories, and research nurses from the wider COG-UK Hospital Onset COVID-19 Infection (HOCI) study. Following debates about what constitutes ‘acceptability’ within healthcare interventions[23], this paper takes an inclusive view of acceptability, and our findings relate to what our participants brought to their interviews rather than using a predefined measure of acceptability.

The HOCI study[24] offered an opportunity to use rapid WGS in real time to improve health outcomes by shaping IPC practice. The HOCI study employed a prospective, non-randomised trial of WGS in 14 acute UK hospital institutes with an integrated qualitative process evaluation. The WGS was trialled to target both rapid (<48 hours) and longer (5-10 days) turnaround times. Steps were taken to minimise the previously identified barriers to sequencing including the development of a bespoke sequencing reporting tool (SRT)[25].

Elsewhere, we report on the specification of the HOCI intervention[26] and provide an in-depth qualitative process evaluation[27] showing how the SRT was used and the ways in which it impacted on IPC and other hospital processes and systems. Whilst we conducted these planned analyses, we were aware of how the diverse participants also talked of issues beyond the particular focus of the HOCI study itself. The current paper reports on this wider material which is broadly concerned with the acceptability of WGS within hospital-based IPC practice, particularly in the context of SARS-CoV-2.

### Research questions

RQ1: What do varied professionals involved in using rapid WGS within hospitals in the COVID-19 pandemic think about its role in managing nosocomial infection?

RQ2: What can be said about HCPs acceptability of rapid WGS for assisting infection prevention and control?

## Methods

### Sampling

From 14 total study sites, a purposive sample of five focal sites were selected for in-depth data collection. Sites were selected to be varied in relation to case rates, hospital-size, familiarity with sequencing and geography. Data collection started after rapid WGS had been used for 14 days.

### Recruitment

Within each site and liaising with a senior member of staff involved in the study, a broad range of professionals involved in the use and implementation of WGS were approached. Those who expressed interest in participating were sent participant information sheets and invited to discuss the project. A mutually convenient time was arranged, and interviews were conducted using online meeting platforms. A single member of the research team (FM) conducted all interviews.

### Participants

Within each of the five selected sites, a sample of between six and nine participants took part. The final sample comprised of 39 participants (n=9 site 1; n = 7 site 2; n=8 site 3; n=8 site 4; n=6 site 5), 27 identified as female (69%) and 12 as males (31%), with an age range of 20-70 (*M*=42). Participants’ roles within the study varied (e.g., clinical fellow, sequencing lab manager, bioinformatician and research nurse).

### Data collection

Sites were enrolled in the study between October 2020 and April 2021. Data collection occurred between December 23rd 2020 and June 2nd 2021 across the peak and decline of the alpha variant. A topic guide was used exploring participants’ experiences of the study in one-to-one interviews (30-90 minutes). Interviews were audio recorded, transcribed by a professional transcribing company and anonymised. Data related primarily to the use rapid WGS as interviews took place following during and after the use of rapid WGS.

### Analysis

Analysis was an iterative process involving both deductive and inductive thematic analysis[28]. First, a research team (n=5) engaged in multiple readings and discussions of the data. After this, an initial coding frame was developed. This contained broad categories of data, some of which were pre-specified and others that were identified in the data (i.e., acceptability of sequencing). Responsible for one site each, the five researchers coded data using the coding-frame, utilising quotes multiple times where necessary. The data from each of the sites were then collated and further inductive analyses were conducted (PF and JM for the current analysis). These inductive analyses were audited closely by at least one other analyst and discussed across the whole qualitative team, in addition to iterative discussions with the wider interdisciplinary team. In the current paper, only analysis relating to the acceptability of sequencing is reported. Given the purposive heterogeneous sample of sites - no attempt was made to compare data collected from sites which actually achieved rapid WGS within the trial versus those that did not, and no attempt was made to situate the analysis in relation to the phase of WGS use as all interviews took place after or during the rapid phase (e.g. rapid vs longer).

### Ethical approval

Ethical approval was given by Cambridge South Research Ethics Committee (20/EE/0118).

## Results

### RQ1: *What do varied professionals involved in using rapid WGS within hospitals in the COVID-19 pandemic think about its role in managing nosocomial infection?*

Inductive thematic analysis yielded three themes, each with subthemes (see Figure 1).

**Figure 1.**
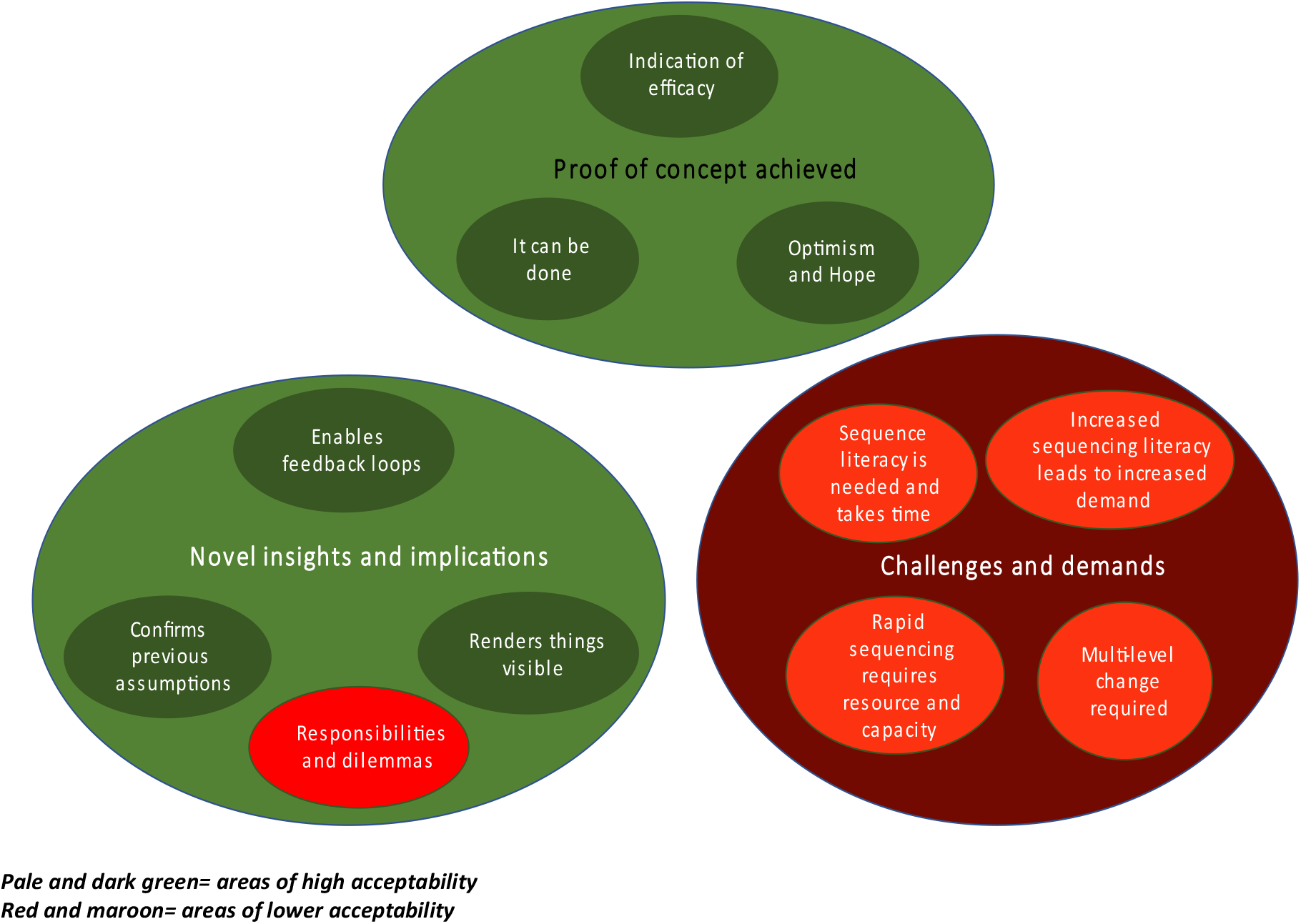
Visualisation of the thematic analysis of acceptability of WGS.

### 1: Proof of concept achieved: the effectiveness and future of rapid sequencing

Analysis suggested, in principle and in practice, the potential of WGS for assisting with HCAIs.

#### a) Despite a pandemic - indications of efficacy

Within the challenging context of the COVID-19 pandemic, most participants shared the view that a significant demonstration of the potential of rapid WGS to aid in managing outbreaks had occurred. Despite, and because of, the pandemic context, sequencing SARS-CoV-2 as part of the HOCI study had delivered benefits. COVID-19 was important as it drove the use of sequencing to address an active problem, allowing IPC teams to learn “*right here, right now” (Site 1, Clinical Academic)*.

> *I think we’ve had some really interesting sequencing results that you know, have been useful both clinically and but certainly extremely useful and interesting in terms of trying to track infections through the Trust and identify sources of infection in the Trust, so from that perspective, you know, it has been useful, and possible, with Covid. (Site 2, Consultant Microbiologist)*

#### b) We have shown it can be done

Participants also believed that many of the longstanding obstacles to implementing real time sequencing had rapidly been overcome (e.g., torpid NHS systems, institutional inertia, expense, risk aversion to innovate).

> *I think the more times things like this is done, the better they’ll get. It’s one of those things where you can either talk about it in theory and tend to moan about the challenges involved, “Well, it’s going to take us, you know, will we ever get the turnaround times low enough”? “No, it doesn’t seem feasible, there’s too much inertia”… we can just do them and see how it goes, evaluate and then look at the next one and improve it. And I think that’s going to be the way forward, I mean, genomics has come of age now and it makes sense to include that in a clinical setting. (Site 4, Bioinformatician)*

Overall, over time, rapid sequencing did deliver: *‘I think it’s been a really powerful tool to be honest’. (Site 3, IPC team member)*.

#### c) Optimism and hope

Many participants were passionate about their sequencing experiences. They felt optimistic about its future potential to improve the management of HCAIs ‘*this looks to me to be very, very important in that we can identify risks, and try to stop problems’ (Site 1, Clinical Academic)*. It was suggested that this may eventually extend, for example, to immediate point of care testing as sustainable, routine practice.

> *It will be interesting to see where it goes in the future, whether there’s anything we can do kind of on the spot, you know, a bit like a blood sugar machine or something. (Site 3, IPC)*

Participants also imagined that rapid sequencing specifically could be used in novel ways; during admission to allocate beds in bays or side rooms; to stratify patients by pathogen strain thus informing and optimising infection control practices and reducing onwards transmission. Participants also talked about the wide-ranging future applicability of sequencing in relation to a span of pathogens (e.g., a range of viral, fungal and bacterial pathogens, including addressing antimicrobial resistance).

### 2: Novel insights and implications of rapid sequencing

Sequencing was seen as an additional tool to existing professional acumen and practice and not seen as a panacea to all IPC problems related to HCAIs. It gave novel insights to be integrated with current practice.

#### a) Sequencing enables feedback loops

Rapid sequencing was seen as pivotal in enabling hospitals to understand more about their own HCAI management such as COVID-19 *‘you appreciate what you’re doing once you see the sequencing and understanding it’ (Site 3, Research Nurse)*. General insights could be gleaned into patient and staff movement or over-reliance on bank staff.

> *It was super-useful to bring stuff back to the clinical teams and show people the patterns for instance, particularly with healthcare workers as well because there’s a lot of fear in the healthcare workers about catching from patients and the use of PPE and obviously we had all that crazy time where we didn’t have the PPE. And I think it definitely helped to be able to give them some real-time information, even if it took a little bit longer than it does now and we kind of brought stuff to the outbreak so we could actually show them and map what had gone on and obviously keep people safe as well. (Site 3, IPC)*

The ability of sequencing to provide feedback on on-going or novel actions was also seen as helping with staff concerns about the efficacy of their own actions, *‘it enables a different level of assurance; conversations as well […*.*] and we had cases, but actually they were quite distinct from each other’ (Site 5, IPC Lead)*. Participants also outlined how using sequencing could be used systematically in the future to evaluate IPC interventions ‘*going forward to show people that actually this doesn’t work or this does work, you know’ (Site 3, IPC)*. ‘*You’ll be able to see that actually there are interventions that you can introduce that would ideally reduce further transmission’ (Site 5, Lead IPC)*

#### b) Rapid sequencing renders things visible in new ways

Sequencing brought particular clarity to understanding hospitals, outbreaks and transmission ‘*it really opened our eyes’ (Site 3, IPC)*: insights concerning new variants; insights into generic transmission routes; the distinction between nosocomial and community acquired infection; the granularity of detail (e.g., ‘*not so sledgehammer’ (Site 1, IPC Lead)*) about the location of nosocomial transmission; or incremental insights into the running of the hospital itself. Throughout, participants talked of sequencing bringing new clarity, and enhanced resolution, to narrow IPC focus.

> *it felt very misty and there were lots of pieces that, I always feel like sometimes with a lot of outbreaks, you feel like there’s loads of pieces up in the air and you’re kind of collecting them all together and ordering them. And this was just massive piece of the jigsaw that initially was missing that we couldn’t actually see who was involved, how far it had gone, if people transferred in were involved, you know, all of that that actually became very clear and it was almost just like, you know, I don’t know, being at the optician and you can suddenly see properly. You’re actually like, I can now see where we’re headed and what’s happened behind us, but also in front of us. (Site 3, IPC)*

New knowledge and insights emerged from rapid sequencing and long-standing assumptions could be questioned ‘*because the sequence doesn’t lie’ (Site 3, Research Nurse)*. It was possible to track the path of variants ‘*it was quite exciting to see it, to see it in real time all the lineages that we would, we had in the hospital (Site 3, Sequencing Team)*. Equally, given the new knowledge sequencing generates, it was possible to assess distinctions between the reality of outbreaks versus assumed ones, ‘*an illusion of an outbreak’ (Site 4, IPC)*.

#### c) Sequencing can confirm previous assumptions and the value of actions already initiated

Rapid sequencing added value to typical IPC practice, bolstering existing approaches to nosocomial infection. It was often used to confirm the assumptions underpinning actions initiated prior to sequencing. This was not surprising given that sequencing is always necessarily retrospective, however rapidly it can be conducted.

> *Yeah, so I think that the confirmation of outbreaks as we’ve had, certainly has helped to drive those kind of interventions [global hospital policies, e*.*g*., *PPE supplies] in places, I think, you know, you’ve got multiple cases in an area, and without the sequencing you’re not sure whether actually you’ve got patient to staff transmission, staff to patient transmission, staff to staff transmission, what’s, you know, you’re not sure what’s going on, whether you’ve got multiple introductions…*.*having sequence confirmation that it’s highly likely that there’s been transmission in an area, does certainly help guide those decisions*.*” (Site 2, Consultant Microbiologist)*

#### d) Responsibility and dilemmas associated with the new knowledge -a double edged sword

Alongside valuable new insights, a few participants highlighted how sequencing led to new dilemmas and responsibilities. Concerns mainly focussed on issues such as blame, potential legal repercussions and ways of mitigating these harms.

> *it’s a difficult one, because I think we do need to understand the role of healthcare workers, and like in any outbreak, so, you know, we deal with Group A strep outbreaks and things or, say, a surgeon who carries MRSA and, you know, you get an increased rate. So it’s a sensitive area but it’s not one we should ever shy away from, to solve a problem you need to understand it. I do think we need very robust planning around protecting healthcare workers’ anonymity in this. And also, moving the whole narrative away from any sort of blame […*.*]. So you’ve got the guilt people feel of having given it to a patient, but equally you’ve got the organisational responsibility for healthcare workers getting it from patients and that comes under, you know, health and safety regulations and all of that (Site 1, Consultant Clinical Microbiologist)*

### 3. Challenges and demands of rapid sequencing

Many participants also talked about multifaceted challenges and demands associated with sequencing.

#### a) Sequence literacy is needed and takes time to embed

Participants outlined the need for people to understand sequencing, what it is doing and what it can do (i.e. sequence literacy). For many professionals the HOCI study was their first experience using sequencing as a practical tool to shape IPC activities, ‘*…so the nurses’ knowledge at the start, it was ‘What is genome sequencing? We have no idea what it is!’’ (Site 5, Bioinformatician)*. Across many sites, a period of learning and gradual engagement culminating in a realisation of what the sequencing was uniquely achieving was reported.

> *I didn’t know how important this study was until I actually understood what information was being given back to us (Site 5, Research Nurse)*.
>
> *I’m not sure clinical staff fully understand why HOCI’s being done. I think that’s changing gradually, I think previously I’m not sure about people knowing the value of it, but I think there might’ve been an underlying worry potentially from clinical staff that the purpose of HOCI might have been to, well why are we getting so many hospital onset Covid admissions, what are staff doing wrong. Not being verbalised but they have initially got a general suspiciousness of us initially, so I do think that has changed (Site 1, Research Nurse)*.

A few participants hinted at the depth of understanding required to use sequencing insights effectively. The usefulness of sequencing depended on its interpretation and a deep understanding of the local context.

#### b) Increased sequence literacy leads to increased demand

Over time, as sequence literacy increased and people came to appreciate the value of sequencing, demand for it increased. Sequencing generated a positive reinforcing feedback loop. In this way, the acceptability and viability of sequencing as a future tool for managing HCAIs and directing IPC was clearly established.

> *they [IPC] definitely became interested and especially as it became clearer and clearer that data, we were generating was useful and timely, yeah, they definitely became very keen and involved (Site 3, Clinical Fellow)*

#### c) Rapid sequencing for IPC requires resource and capacity

The more rapid WGS is, the better it is at directing IPC to affect preventable nosocomial infection. Substantial ‘start-up costs’ were essential for the practical application of rapid sequencing within some hospitals. Across sites, experience of sequencing was noticeably heterogeneous; some sites were readied to capitalise on sequencing capacity more than others. Some sites, for example, had both on-site diagnostic and sequencing labs; others had to forge new relationships with academic institutions or other sites to attain this capacity. These initial ‘costs’ were keenly felt by some. Basic requirements included means of rapidly transporting samples, timely access to diagnostic and sequencing labs, the availability of bioinformatician expertise, capacity to collate necessary meta-data and training for a variety of staff.

> *I would put resource into ensuring that there were sort of medical or senior nursing staffing input who understand what sequence interpretation and infection control, so I think that person, so I think some training actually within infection control about how to interpret data and so on would be really, really useful. I think you need significant sort of resource going into, I think it would be better to place the sequencing in the NHS and to resource the NHS to do the sequencing and then, but that’s another step. (Site 1, Clinical Academic)*

A minority of participants also talked about the longer-term costs associated with providing a sustainable rapid sequencing service. Given rapidity was associated with usefulness they questioned at which level (e.g., government) resources should be provided.

#### d) Rapid sequencing for IPC requires multi-levelled change

Participants also outlined that beyond initial ‘basic’ re uirements, further changes in the ways technologies, people, teams and institutions are connected must occur to get the most out of sequencing. The function of rapid sequencing (and degrees of its usefulness) depended on effectively consolidating these wider assets. The degree to which the co-ordination of resources can come together ma es the difference between ‘*informing the onward management’ (Site 4, Surveillance Officer)* of an outbreak versus being ‘*an academic exercise, a retrospective review and it’s not timely enough to have an impact’ (Site 4, Surveillance Officer)*, or more positively, ‘*more slow burning strategic interventions’ (Site 2, Consultant Microbiologist)*.

> *It’s timeliness and confidence that you’ve got; you’ve covered everything. If you don’t have those two things with sequencing, then it can’t really change your decisions that much, to be absolutely honest (Site 4, IPC)*.

### RQ2: What can be said about HCPs acceptability of rapid WGS for assisting infection prevention and control?

WGS was almost universally accepted by the professionals who took part in this research. In relation to seven key conceptualisations of acceptability[^23]^, our findings (Figure 1) support the ‘affective dimensions’ of WGS and detail its ‘perceived effectiveness’. They suggest that issues of WGS ‘burden’, ‘coherence’ and ‘self-efficacy’ are particularly important in the early days of implementing WGS but their importance reduces over time. Our analysis also spea s to acceptability in relation to ‘ethicality’ and a few of our participants highlighted the dilemmas and responsibilities that WGS elicited. Finally, our analysis leaves acceptability - questions of ‘opportunity costs’ - relatively unaddressed.

## Discussion

This paper provides novel insights into acceptability of rapid WGS as an applied tool used to direct IPC and reduce nosocomial infections in the context of the COVID-19 pandemic. To our knowledge, it is the first, globally, to detail the perspectives of a wide-ranging group of professionals (from bioinformaticians to IPC nurses) who used sequencing in real time with the aim to reduce HCAIs. Participants shared their rich, detailed experiences and our thematic analysis structured their accounts to highlight three main aspects of acceptability (i.e., ‘*Proof of concept achieved’*; ‘*Novel insights and implications’*; ‘*Challenges and demands*). The analysis suggests the acceptability of rapid WGS as an additional IPC tool among HCPs, albeit with multi-facetted challenges. Critically, however, many of the demands were short-lived and associated with the *novelty* of implementing sequencing as a practical IPC tool. The context of the COVID-19 pandemic was also important; it drove the HOCI study, releasing research funding to systematically reduce long-standing barriers, yet also compromised it with the impacts of the alpha variant overwhelming IPC ability to act on sequencing insights and making sequence reports more challenging to interpret and less discriminatory due to significant sequence homogeneity.

### Strengths and limitations

The key strengths of our work are centred on the effective use of inductive thematic analysis (i.e., learning from the participants and their data) to instigate the paper - although not pre-planned, the varied participants all talked at length about acceptability, meriting this analysis. Other strengths include the use of multiple qualitative analysts and an iterative process of working with the larger interdisciplinary team. This use of multiple perspectives ensures a stronger and more accessible analysis.

Limitations relate to the potential for a positive bias towards acceptability and perceived usefulness of WGS for HCAI, given the involvement and on-going dialogue of participating sites with the wider HOCI research team and that measures had been taken to minimise the previously identified barriers to sequencing. Any potential bias may well have been amplified by the sole use of a qualitative design (rather than mixed methods to triangulate findings and assess wider applicability). Further limitations include the limited number of sites in which data collection took place (five of fourteen sites). It should be noted however that an interactive day-long event including representation from all participating sites provided broad agreement with these qualitative insights.

In translating our findings, it is also important to acknowledge that data collection took place following emergence of the alpha variant of SARS-CoV-2 in the UK[29]. This led to a sharp increase in the community incidence rates of SARS-CoV-2, and in the associated health burden and rates of nosocomial transmission within hospitals. Data were therefore collected in a particularly challenging context and time within the COVID pandemic especially for sites implementing sequencing for the first time.

### Implications and future research

Our analysis suggests that rapid-sequencing to assist with managing HCAIs is perceived to be acceptable - at least within the COVID-19 pandemic context. Largely positive perspectives on rapid sequencing for HCAIs were highlighted - although several key challenges were also detailed (Figure 1). To some extent, these challenges may have related to the novelty of sequencing in some sites and, across all sites, the teething troubles of developing a functioning sequencing pathway with a rapid turnaround time (i.e., ensuring sequencing data could directly affect IPC actions). Further research is needed to explore the future of sequencing for managing HCAIs as it is currently unclear if we can capitalise on recent progress in implementing routine sequencing to build sequencing capacity for the future. It is unknown, for example, if the extent to which start-up costs for implementing a sequencing pathway are indeed one-off or if the largely positive findings reported here can translate to other contexts; beyond the COVID-19 pandemic; in epidemic and endemic scenarios; across a wider range of pathogens (including those that are less lethal); across diverse settings (e.g., care homes). As such, important and interdisciplinary research questions remain that address the balance of barriers to implementation versus benefit over time.

## Conclusion

Our qualitative analysis has highlighted the acceptability and promise of rapid sequencing for HCAIs among HCPs within the context of the COVID-19 pandemic, from a small sample of UK NHS hospitals taking part in a multisite study. It shows that, even under extremely challenging conditions, rapid sequencing can deliver useful insights, and, with further work, such rapid sequencing could be transformative in relation to HCAI management, potentially representing a step change for IPC.

## Data Availability

Data is not available given it is qualitative and anonymity of participants must be protected

## Acknowledgements

We would like to particularly acknowledge the support of NHS Greater Glasgow and Clyde Clinical Research Facility.

We also acknowledge the support of the independent members of the Joint Trial Steering Committee and Data Monitoring Committee (TSC-DMC): Prof Marion Koopmans (Erasmus MC), Prof Walter Zingg (University of Geneva), Prof Colm Bergin (Trinity College Dublin), Prof Karla Hemming (University of Birmingham), Prof Katherine Fielding (LSHTM). As well as TSC-DMC non-independent members: Prof Nick Lemoine (NIHR CRN), Prof Sharon Peacock (COG-UK). We would also thank members of COG-UK who have directly supported the study: Dr Ewan Harrison (Cambridge University), Dr Katerina Galai (PHE), Dr Francesc Coll (LSHTM), Dr Michael Chapman (HDR-UK), Prof Thomas Connor and team (Cardiff University), Prof Nick Loman and team (University of Birmingham). We also thank the COG-UK Consortium, and the UK National Institute for Health Research Clinical Research Network (NIHR CRN).

## Conflict of interest statement

Paul Flowers – <u>No competing interests declared</u>

Julie McLeod-<u>No competing interests declared</u>

Fiona Mapp-<u>No competing interests declared</u>

Oliver Stirrup <u>No competing interests declared</u>

James Blackstone <u>No competing interests declared</u>

Luke Snell <u>No competing interests declared</u>

Christine Peters <u>No competing interests declared</u>

Emma Thomson, <u>No competing interests declared</u>

Alison Holmes <u>No competing interests declared</u>

James Price <u>No competing interests declared</u>

Dave Partridge <u>No competing interests declared</u>

Laura Shallcross <u>No competing interests declared</u>

Thushan I de Silva <u>No competing interests declared</u>

Judith Breuer <u>No competing interests declared</u>

## Funding statement

This work was supported by funding from the Medical Research Council (MRC) part of UK Research & Innovation (UKRI), the National Institute of Health Research (NIHR) [grant code: MC_PC_19027], and Genome Research Limited, operating as the Wellcome Sanger Institute.

## Notes

### Competing Interest Statement

The authors have declared no competing interest.

### Author Declarations

Ethical approval was given by Cambridge South Research Ethics Committee (20/EE/0118).

